# Meta-analytical evidence on mental disorder symptoms during the COVID-19 pandemic in Latin America

**DOI:** 10.1101/2021.06.21.21259299

**Authors:** Stephen X. Zhang, Kavita Batra, Tao Liu, Rebecca Kechen Dong, Wen Xu, Allen Yin, Andrew Delios, Bryan Z. Chen, Richard Z. Chen, Saylor Miller, Xue Wan, Jiyao Chen

**Affiliations:** Faculty of Professions, University of Adelaide, Adelaide, Australia; Office of Research, School of Medicine, University of Nevada, Las Vegas NV 89102; College of Economics and Management, Southwest University, China; Business School, University of South Australia, Adelaide SA5001, Australia; Nottingham University Business School China, University of Nottingham Ningbo China, Ningbo 315100, China; School of Humanities, Southeast University, Nanjing, China; University of Adelaide, SA 5001, Australia; Crescent Valley High School Crescent Valley High School, Corvallis, OR 97330, USA; Crescent Valley High School, Corvallis, OR 97330, USA; College of Business, Oregon State University, OR 97330, USA; School of Economics and Management, Tongji University, Shanghai, China; College of Business, Oregon State University, Corvallis OR 97330

**Author notes:** Correspondence to: Jiyao Chen, College of Business, Oregon State University, Corvallis OR 97330.

**Keywords:** Mental health, Prevalence, Healthcare Workers, Anxiety, Depression, Distress, Insomnia

## Abstract

**Objective:** There is a lack of evidence related to the prevalence of mental disorder symptoms as well as their heterogeneities during the COVID-19 pandemic in Latin America, a continent across the equators. The current study aims to provide meta-analytical evidence on mental disorder symptoms during COVID-19 among frontline healthcare workers, general healthcare workers, the general population, and university students in Latin America.

**Methods:** Bibliographical databases, such as *PubMed, Embase, Web of Sciences, PsycINFO*, and *medRxiv*, were systematically searched to identify pertinent studies up to Februry 6, 2021. Two coders performed the screening using predefined eligibility criteria. Studies were assigned quality scores using the Mixed Methods Appraisal Tool. The double data extraction method was used to minimize data entry errors.

**Results:** A total of 33 studies with 101,772 participants in Latin America were identified. The pooled prevalence of anxiety, depression, distress, and insomnia was 32%, 27%, 32%, and 35%, respectively. There was a higher prevalence of mental health symptoms in South America compared to Central America (33% vs. 27%, p <0.001). The pooled prevalence of mental health symptoms in the general population, general healthcare workers, frontline healthcare workers, and students in Latin America was 33%, 31%, 37%, and 36%, respectively.

**Conclusion:** The high yet heterogenous level of prevalence of mental disorder symptoms emphasizes the need for appropriate identification of psychological interventions in Latin America.

## Introduction

The continent of Latin America of 34 countries or territories has had the second amount of cases and deaths per capita for coronavirus disease (COVID-19)(Burki, 2020),(Ríos, 2021; World Health Organization, 2020). Latin America is vulnerable to the destructive outbreak for several reasons^1,4,5^ including long-standing structural and socioeconomic inequities(Burki, 2020; Carvalho et al., 2015; Dávila-Cervantes & Agudelo-Botero, 2019), over 20% of the population in poverty, lack of healthcare access, underfunded healthcare systems, poor governance or political dynamics, and a high burden of chronic and metabolic health conditions, and lack of preparedness to fight the pandemic(Malta et al., 2020)^-8^. Reportedly, there is a considerable increase in psychological morbidities among several demographic groups, including healthcare workers, the general population, and students(Campos et al., 2021). Latin America is also a vast continent, where tropical regions span across almost all Latin American countries, and regional disparities on mental health has been reported(Malta et al., 2020), but we still lack evidence on the prevalence of mental disorder symptoms as well as their heterogeneities in Latin America during the COVID-19 pandemic.

Recently, meta-analyses have provided early global evidence on the prevalence of mental disorder symptoms across groups, including healthcare workers, the general population, and students(Batra et al., 2021; K Batra et al., 2020; X. Chen et al., 2021; M Luo et al., 2020; Pappa et al., 2020). These previous reports included very few studies based on Latin American samples, therefore meta-analytical evidence for Latin America is lacking. With emerging studies on mental health in Latin America, it is critical to synthesize meta-analytical evidence to provide integrated data on mental health among key demographic groups in Latin America during the COVID-19 pandemic. Therefore, this meta-analysis aims to investigate the pooled prevalence of mental disorder symptoms during the COVID-19 pandemic among frontline healthcare workers, general healthcare workers, the general population, and university students in Latin America. We first perform subgroup analysis breaking Latin America based on South America (a majority but not all countries are in tropical regions) and Central America (all countries are entirely tropical).

## Methods

### Protocol Registration

We followed the Preferred Reporting Items for Systematic Reviews and Meta-Analyses (PRISMA) statement 2020 (Liberati et al., 2009) to guide our meta-analysis and registered it with the International Prospective Register of Systematic Reviews (PROSPERO: CRD42020224458).

### Eligibility Criteria

The search targeted observational studies that assessed the prevalence of psycho-morbid symptoms of anxiety, depression, distress, and insomnia among frontline healthcare workers, general healthcare workers, the general population aged 18 years or above, and university students in Latin America. A priori inclusion criteria were established to identify eligible studies that used established psychometric survey tools, used English language, and were available as full-texts. Studies which targeted other populations, including children, adolescents, and certain subgroups (e.g. pregnant women), were excluded. Other study designs, such as reviews and meta-analyses, qualitative, mixed methods, case reports, studies published only as abstracts, biochemical and experimental studies, or articles lacking the use of psychometric robust instruments or with ambiguous methodology to identify prevalence were also excluded. Studies based on non-Latin American countries were excluded. Studies with unclear methodology and results were reviewed carefully and a researcher (WX) of our study attempted to contact authors to seek the information in several instances: 1) if the study reported estimates for both targeted and excluded populations, posing challenges for us to delineate the prevalence rate for the population of interest to our study; 2) if the study did not report the prevalence as proportions; 3) if the study did not specify cut-off sores for levels of severity; or 4) if the study was missing crucial information such as response rate, duration of data collection, and gender distribution.

### Data Sources and Search Strategy

This meta-analysis is part of a large project on meta-analysis of mental health symptoms during COVID-19. Bibliographic databases, such as PubMed, Embase, PsycINFO, and Web of Sciences, were searched on February 6, 2021. MedRxiv was also searched for preprints. Search algorithms specific to each database were used to yield a comprehensive pool of literature. A detailed search strategy appears in Table S1.

### Phases of Screening

A researcher (JC) exported the search results from various databases into Endnote to remove duplicates and then imported them into Rayyan for subsequent screening. Two reviewers (AD & BZC) independently screened the titles and abstracts of all papers in accordance with the prespecified eligibility criteria. The eligible abstracts proceeded to full-text screening for possible inclusion. Any conflicts between reviewers were resolved by a third reviewer (RKD).

### Data Extraction

A codebook was developed for standardization and consistency. The final studies included from the screening process were sent to three groups (two reviewers in each group, WX & AY, BZC & AD, RZC & SM) for thorough investigation and extraction of relevant data elements into a coding book. Standardized codes were used to record pertinent variables, including author, title, country, duration of data collection, study design, population, sample size, response rate, female proportion rate, mean age, psychological outcome, severity level of outcome, type of survey instruments with cut-off scores, and prevalence of psycho-morbid events. The severity of psychological outcomes of interest was coded as above mild, moderate above, and severe levels (if available). The studies that reported only mild, moderate, and severe prevalence data were recoded into mild above, moderate above, and severe prevalence for consistency purposes. The severity levels in studies that only reported the overall prevalence were determined based on cut-off scores (if available). After finishing independent coding, all the extracted data elements were subject to a second round of review by the coders to identify any discrepancies. In case of disagreements, a third reviewer (TL) was asked for the final decision. The third reviewer helped to achieve consensus through re-verification and discussion.

### Risk of Bias (RoB) Assessment

The Mixed Methods Appraisal Tool (MMAT) with seven questions was used as a quality assessment tool(Hong et al., 2018; Pablo et al., 2020; Usher et al., 2020). Two reviewers independently assessed and assigned scores to the studies using the tool dictionary and guidelines. Disagreements were resolved through discussion with the lead reviewer (RKD). The quality scores ranged from 0 to 7 (highest quality). Studies were categorized as high, medium, or low quality, if they attained the score of 6, 5 to 6, or <5 respectively^18^.

### Effect Measure and Data Analysis

Using Version 16.1 of Stata (metaprop package), a random effects model was used to compute the pooled estimates(DerSimonian & Laird, 1986). To quantify heterogeneity, I^*2*^ statistics was used and I^2^ over 75% was considered substantial(Higgins et al., 2019). Visual inspection of the Doi plot and the Luis Furuya–Kanamori(Furuya-Kanamori et al., 2018) index were used to assess publication bias(Kounou et al., 2020; Yitayih et al., 2020). Event ratio was used as the primary effect measure for the pooled estimates.

## Results

### Screening of Studies

A total of 6,949 records were identified through searching bibliographical databases and other sources (Figure 1). After removing 3,603 duplicates, a total of 3,346 records advanced to the phase of screening. After excluding 2,662 records that did not pass the title and abstract screening, only 684 records were identified as eligible for full-text screening. Of the 684 studies, findings of 29 studies were verified through emails with their authors. We received responses for 8 studies, which were included in the final pool. We identified a total of 168 studies of which 135 did not study populations in Latin American countries (Figure 1). Therefore, only 33 studies, focused on populations in Latin America, were used in the final data extraction and analysis.

**Figure 1:**
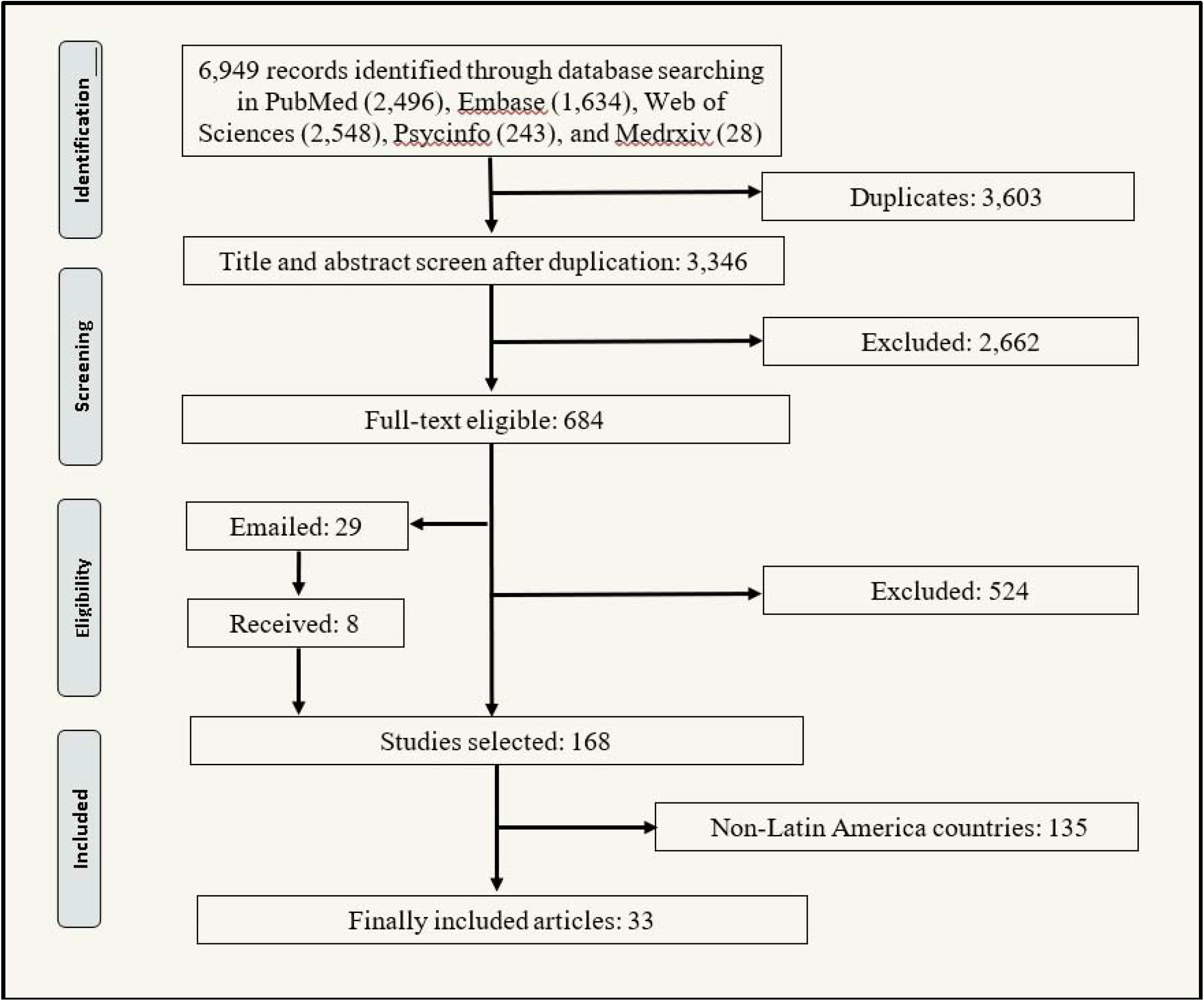
PRISMA diagram detailing the screening process.

## Study Characteristics

A total of 36 unique samples from 33 studies(Badellino et al., 2020; Campos et al., 2020; Cénat et al., 2021; Chen et al., 2020a; Chen et al., 2020b; Civantos et al., 2020; Cortés-Álvarez et al., 2020; Dal’Bosco et al., 2020; De Boni et al., 2020; Fernández et al., 2020; Feter et al., 2021; Giardino et al., 2020; Goularte et al., 2021; Guiroy et al., 2020; Loret de Mola et al., in press; Malgor et al., 2020; Martinez et al., 2020; Medeiros et al., 2020; Monterrosa-Castro et al., 2020; Mora-Magaña et al., 2020; Paz et al., 2020; Puccinelli et al., 2021; Robles et al., 2020; Samaniego et al., 2020; Schuch et al., 2020; Scotta et al., 2021; Serafim et al., 2021; Torrente et al., 2021; Werneck et al., 2021; Jaime A. Yáñez et al., 2020; Jaime A Yáñez et al., 2020; Stephen X Zhang et al., 2021; Stephen Xu Zhang et al., 2021) involving 101,772 participants from Latin America were included in this meta-analysis (Table 1). Some studies include multiple independent samples. For example, one study^47^ examined the prevalence of both general healthcare workers and frontline healthcare workers. Among them, 20 samples (55.5%) were of general populations^3, 6, 8-10, 12, 15, 17-22, 25, 28, 30-32^, 2 samples (5.5%) were of frontline healthcare workers(Dal’Bosco et al., 2020; Robles et al., 2020),12 samples (33.3%) were from general healthcare workers (Chen et al., 2020a; Chen et al., 2020b; Civantos et al., 2020; Giardino et al., 2020; Guiroy et al., 2020; Malgor et al., 2020; Monterrosa-Castro et al., 2020; Mora-Magaña et al., 2020; Robles et al., 2020; Samaniego et al., 2020; Jaime A. Yáñez et al., 2020; Jaime A Yáñez et al., 2020; Stephen X Zhang et al., 2021), and 2 samples (5.5%) were based on university students(Medeiros et al., 2020; Scotta et al., 2021). Of the 33 studies, 17 were from Brazil (Table 1). Except for two (6.1%) longitudinal cohort studies(Feter et al., 2021; Loret de Mola et al., in press), the majority of the studies were cross-sectional (93.9%). The sample size varied from 57 to 43,995 participants. The participation rates varied from 11.4% to 100.0% with a median value of 66.0%. The female proportions among the 36 samples varied from 3.4% to 100% with a median of 69.8%.

**Table 1.** Characteristics of the studies on mental health in Latin America during the COVID-19 pandemic

| Characteristics | Total number of studies/samples* | Percent | Level of analysis |
| --- | --- | --- | --- |
| **Overall** | **33/36** | **100** |  |
| **Outcome #** |  |  | Prevalence |
| Anxiety | 56 | 41.48 |  |
| Depression | 49 | 36.3 |  |
| Distress | 14 | 10.37 |  |
| Insomnia | 16 | 11.85 |  |
| **Severity#** |  |  | Prevalence |
| Above mild | 52 | 38.52 |  |
| Above moderate | 45 | 33.33 |  |
| Above severe | 30 | 22.22 |  |
| Overall | 8 | 5.93 |  |
| **Population** |  |  | Sample |
| Frontline healthcare workers | 2 | 5.56 |  |
| General healthcare workers | 12 | 33.33 |  |
| General population | 20 | 55.56 |  |
| Students | 2 | 5.56 |  |
| **Sampling Country** |  |  | Sample |
| Argentina | 5 | 13.89 |  |
| Bolivia | 1 | 2.78 |  |
| Brazil | 17 | 47.22 |  |
| Colombia | 1 | 2.78 |  |
| Ecuador | 2 | 5.56 |  |
| Haiti | 1 | 2.78 |  |
| Mexico | 5 | 13.89 |  |
| Paraguay | 1 | 2.78 |  |
| Peru | 2 | 5.56 |  |
| Mixed | 1 | 2.78 |  |
| **Quality** |  |  | Study |
| High | 13 | 39.4 |  |
| Medium | 20 | 60.6 |  |
| **Design** |  |  | Study |
| Cohort | 2 | 6.06 |  |
| Cross-sectional | 31 | 93.94 |  |
| **Publication** |  |  | Study |
| Preprint | 2 | 6.06 |  |
| Published | 31 | 93.94 |  |
| **Overall** | **Mean (median)** | **Range** |  |
| Number of participants | 2827 (615) | 57 – 43,995 | Sample |
| Female proportion | 65.1% (69.8%) | 3.4% – 100% | Sample |
| Response rate | 63.3% (66.0%) | 11.4% – 100% | Sample |

\*Some studies include multiple independent samples. For example, one study<sup>47</sup> examined the prevalence of both general healthcare workers and frontline healthcare workers. #One independent sample in a study may report anxiety, depression, and insomnia at the levels of mild above, moderate above, and severe. Therefore, the total number of prevalence is larger than the total number of independent samples.

### Estimates of Pooled Prevalence of Psychological Morbidity Symptoms

In Latin America, 32 samples from 29 studies(Badellino et al., 2020; Campos et al., 2020; Cénat et al., 2021; Chen et al., 2020b; Civantos et al., 2020; Cortés-Álvarez et al., 2020; Dal’Bosco et al., 2020; De Boni et al., 2020; Fernández et al., 2020; Feter et al., 2021; Giardino et al., 2020; Goularte et al., 2021; Loret de Mola et al., in press; Malgor et al., 2020; Martinez et al., 2020; Medeiros et al., 2020; Monterrosa-Castro et al., 2020; Mora-Magaña et al., 2020; Paz et al., 2020; Puccinelli et al., 2021; Robles et al., 2020; Samaniego et al., 2020; Schuch et al., 2020; Serafim et al., 2021; Torrente et al., 2021; Werneck et al., 2021; Jaime A. Yáñez et al., 2020; Stephen X Zhang et al., 2021; Zhang et al., in press) reported the prevalence of anxiety symptoms among 99,064 participants. Among all the anxiety survey tools used, the Generalized Anxiety Symptoms 7-items scale (GAD-7) was the most common (51.7%), followed by the Hospital Anxiety and Depression Scale (HADS) (13.8%), Depression, Anxiety and Stress Scale – 21 Items (DASS-21) (10.3%), and Brief Symptom Inventory 18 (BSI-18) (3.5%). The cut-off values to determine the overall prevalence as well as the severity of anxiety varied across studies. In the random-effects model, the pooled prevalence of anxiety was 32% (95% CI: 27%–38%, I^2^ = 99.8%) in the 29 studies (Figure 2A). A total of 27 samples from 24 studies(Badellino et al., 2020; Campos et al., 2020; Civantos et al., 2020; Cortés-Álvarez et al., 2020; Dal’Bosco et al., 2020; De Boni et al., 2020; Fernández et al., 2020; Feter et al., 2021; Giardino et al., 2020; Goularte et al., 2021; Guiroy et al., 2020; Loret de Mola et al., in press; Martinez et al., 2020; Medeiros et al., 2020; Mora-Magaña et al., 2020; Paz et al., 2020; Puccinelli et al., 2021; Robles et al., 2020; Samaniego et al., 2020; Schuch et al., 2020; Serafim et al., 2021; Torrente et al., 2021; Stephen X Zhang et al., 2021; Zhang et al., in press) reported the prevalence of depression among 53,622 respondents. Among all the depression survey tools, the Patient Health Questionnaire (PHQ)-9 was the most frequently used (45.83%), followed by HADS (16.67%), DASS-21 (12.5%), and Beck Depression Inventory (BDI) (4.17%). On analysing the random-effects model, the pooled prevalence of depression was 27% (95% CI: 27%–38%, I^2^ = 99.8%) among the 24 studies (Figure 2B). Eight studies(Chen et al., 2020a; Civantos et al., 2020; Cortés-Álvarez et al., 2020; Fernández et al., 2020; Reidy, 2020; Samaniego et al., 2020; Jaime A. Yáñez et al., 2020; Stephen Xu Zhang et al., 2021) studied mental distress among 7,644 participants. In the random-effects model, the pooled prevalence of distress was 32% (95% CI: 22%–42%, I^2^ = 99.9%) (Figure 2C). Seven samples of the five studies(Giardino et al., 2020; Goularte et al., 2021; Robles et al., 2020; Samaniego et al., 2020; Scotta et al., 2021) studied insomnia among 11,088 respondents. The Insomnia Severity Index (ISI) was used most often to measure insomnia followed by DSM. In the random-effects model, the pooled prevalence of insomnia was 35% (95% CI: 24%–48%, I^2^ = 99.6%) (Figure 2D).

**Figure 2A.**
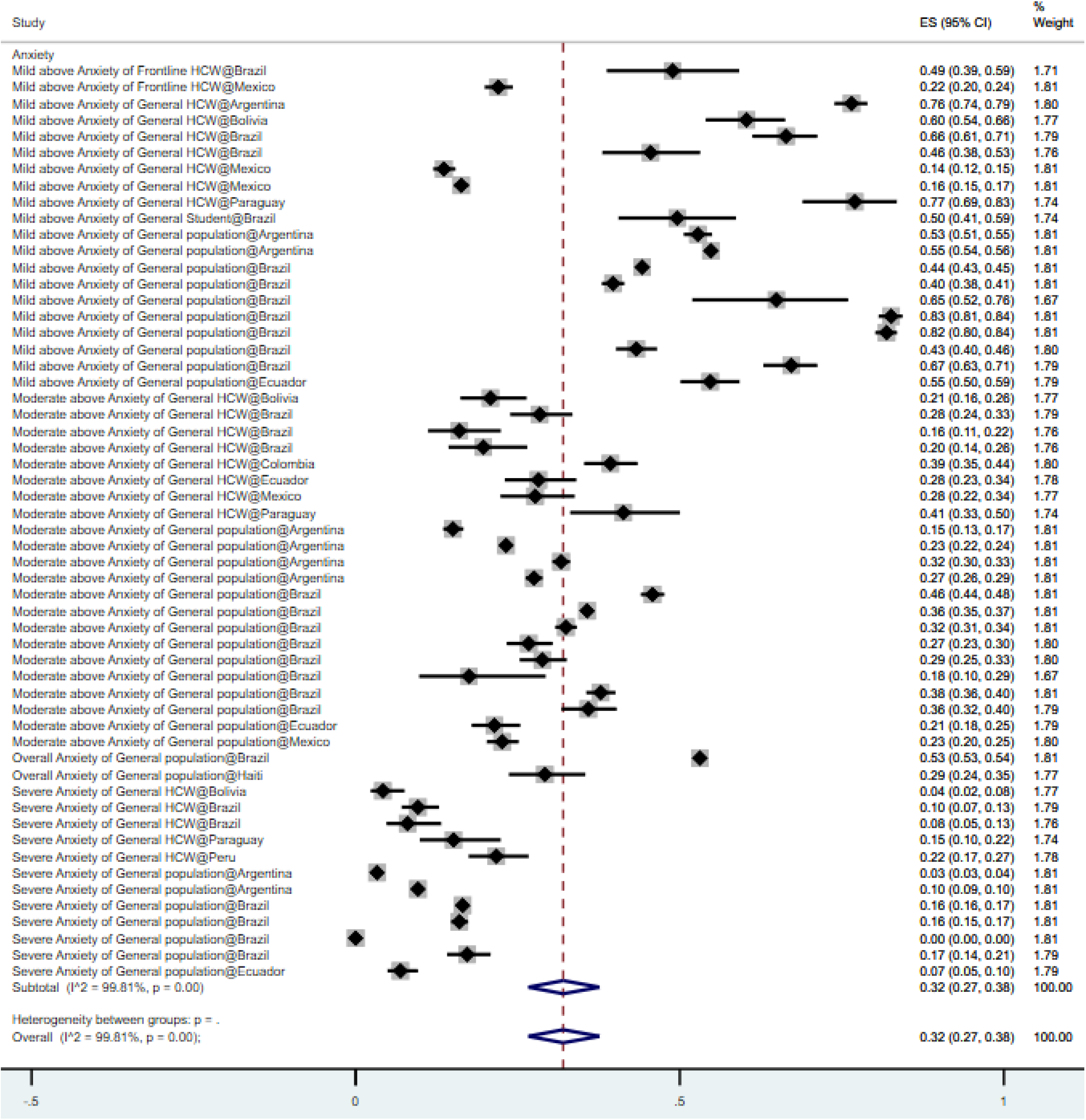
Forest plot indicating the pooled prevalence of anxiety among included studies. The square markers indicate the prevalence of anxiety symptoms among population groups of interest. The diamonds represent the pooled estimates.

**Figure 2B.**
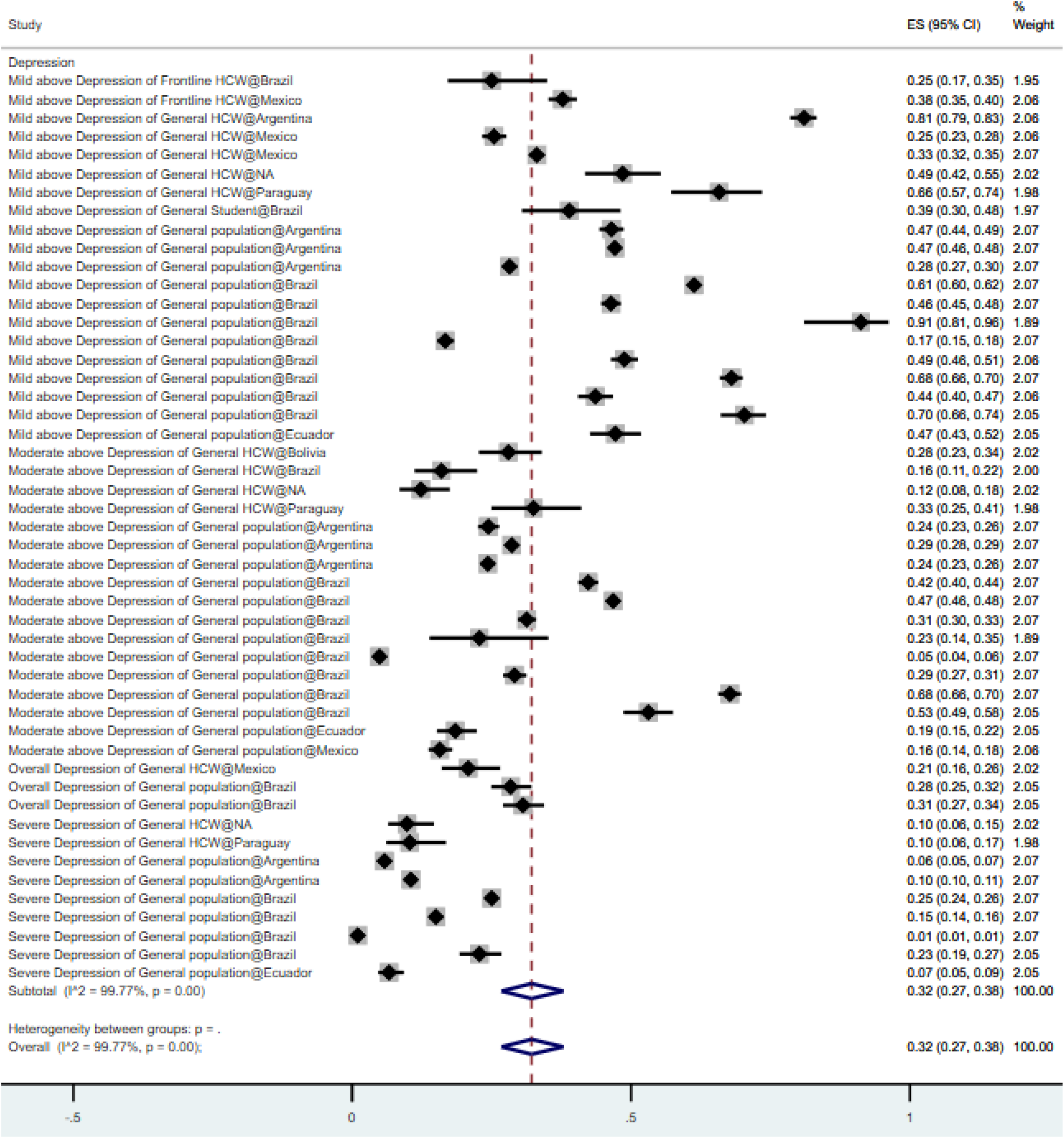
Forest plot indicating the pooled prevalence of depression among included studies. The square markers indicate the prevalence of depression symptoms among population groups of interest. The diamond represents the pooled estimates. NA = Not applicable, when study location was not specified.

**Figure 2C.**
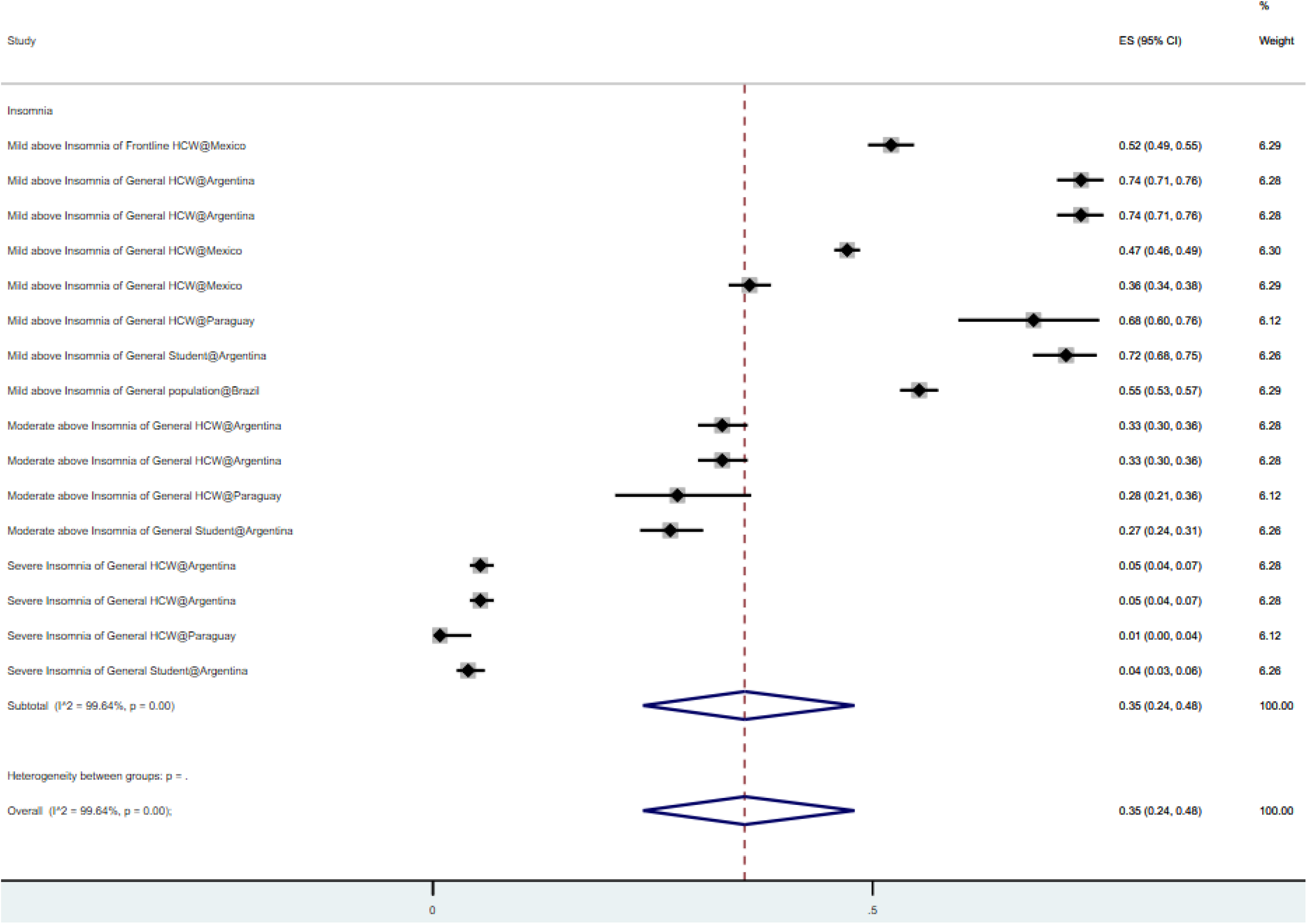
Forest plot indicating the pooled prevalence of distress among included studies. The square markers indicate the prevalence of distress symptoms among population groups of interest. The diamond represents the pooled estimates.

**Figure 2D.**
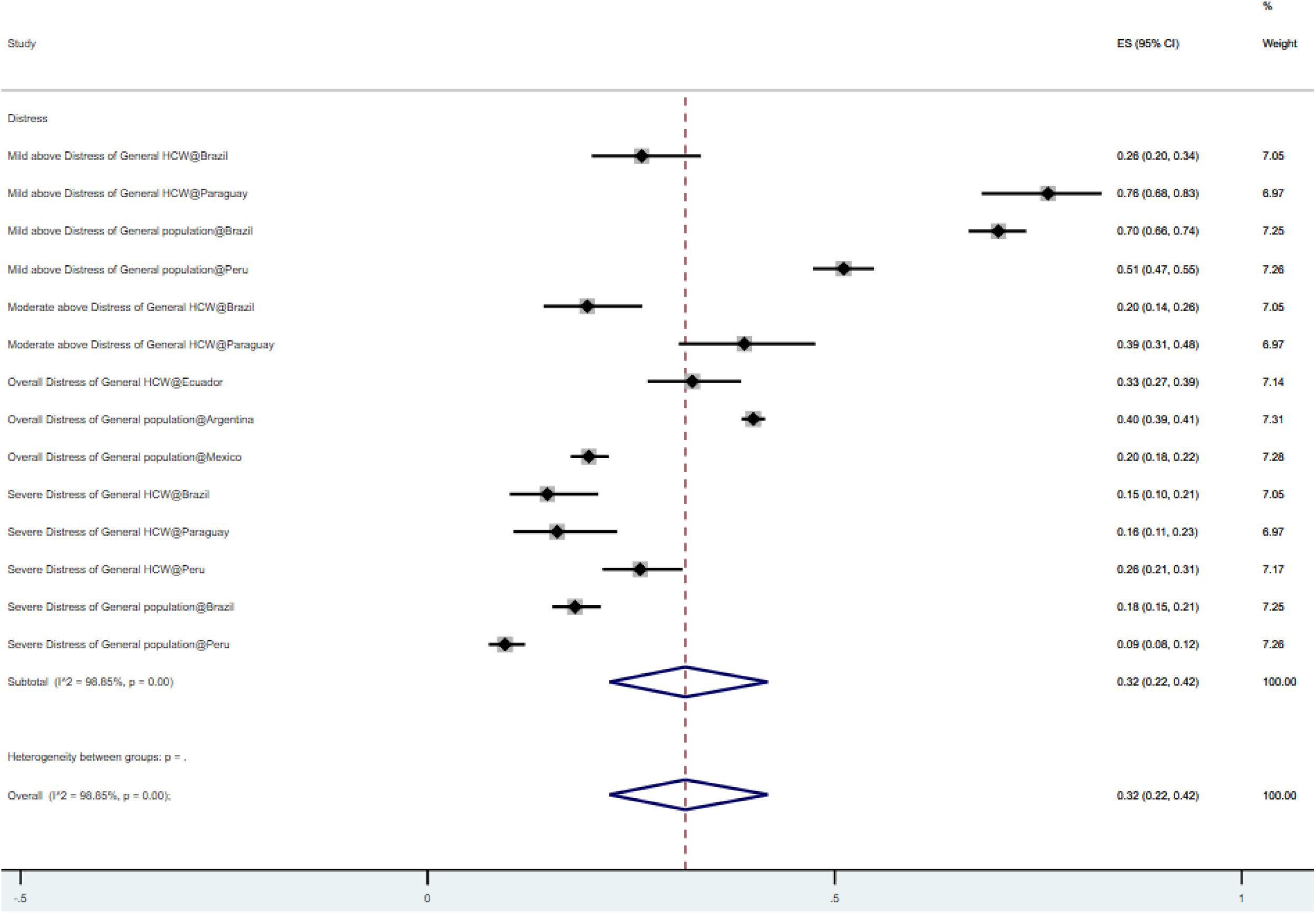
Forest plot indicating the pooled prevalence of insomnia among included studies. The square markers indicate the prevalence of insomnia symptoms among population groups of interest. The diamond represents the pooled estimates.

The overall prevalence of mental health symptoms in frontline healthcare workers, general healthcare workers, the general population, and students in Latin America was 37%, 31%, 33%, and 36%, respectively. The overall prevalence rates of mental health symptoms that exceeded the cut-off values of mild, moderate, and severe symptoms were 53%, 28%, and 10%, respectively (Table 2). The pooled prevalence rates of mental health symptoms in South America, Central America, countries speaking Spanish, and countries speaking Portuguese were 33%, 27%, 30%, and 36%, respectively (Table 2).

**Table 2.**
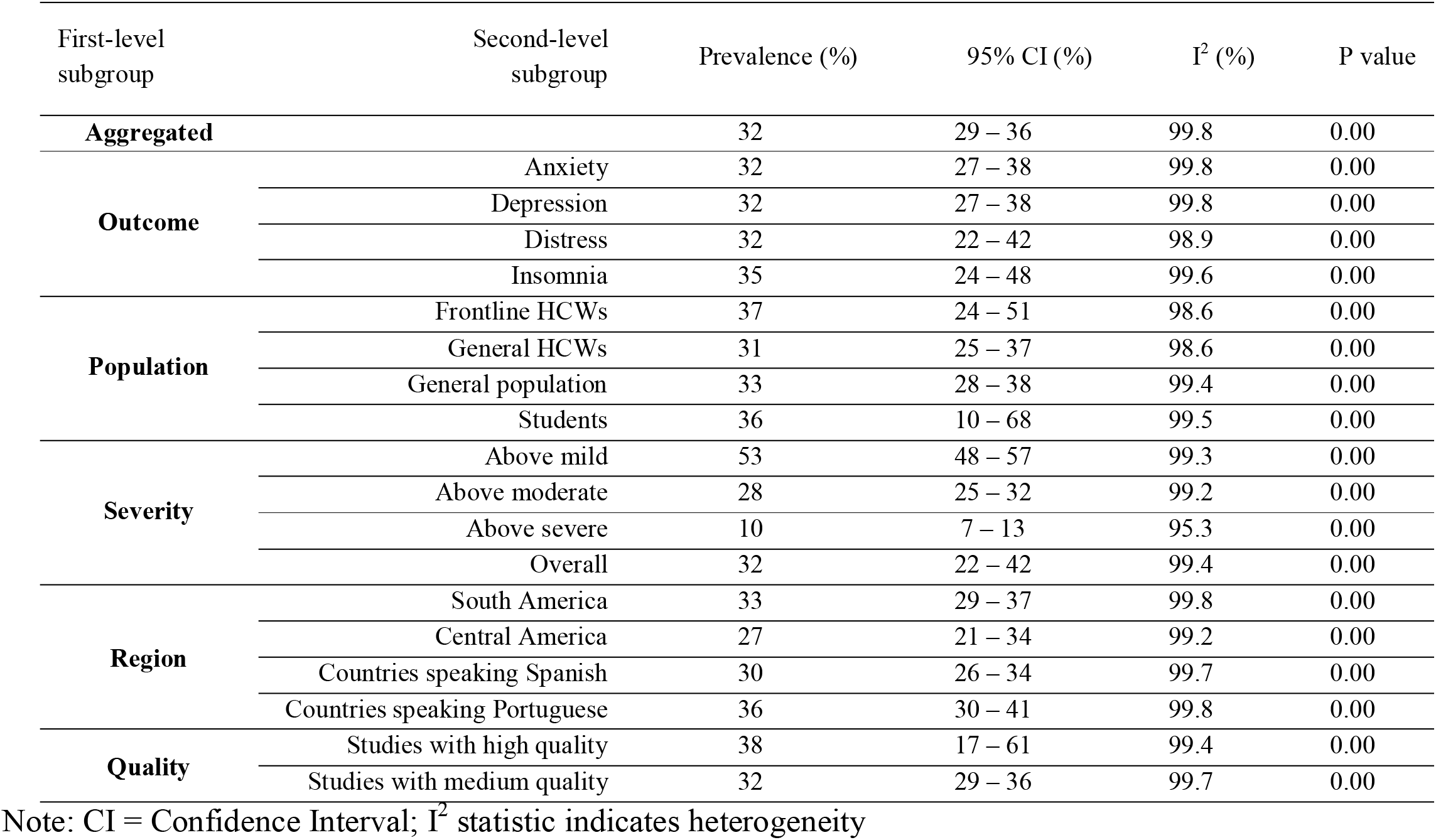
Pooled prevalence estimates of mental health symptoms by outcome, population, severity and region subgroups during the COVID-19 pandemic

### Quality of the Studies

Of all studies, 13 studies (39.4%) were of high quality (score of 7), and 20 studies (60.6%) were of medium quality^18^ (Table 1). There were no studies of low quality. The subgroup analysis suggests the studies with high quality reported lower prevalence of mental health issues in Latin America (Table 2).

### Detection of Publication Bias

The Doi plot and Luis Furuya–Kanamori index were used to quantify publication bias rather than the funnel plot and Egger’s statistics(Furuya-Kanamori et al., 2018; Kounou et al., 2020). The symmetrical, hill-shaped Doi plot and a Luis Furuya–Kanamori (LFK) index of -0.81 indicated ‘no asymmetry’ and a lower likelihood of publication bias (Figure 3).

**Figure 3.**
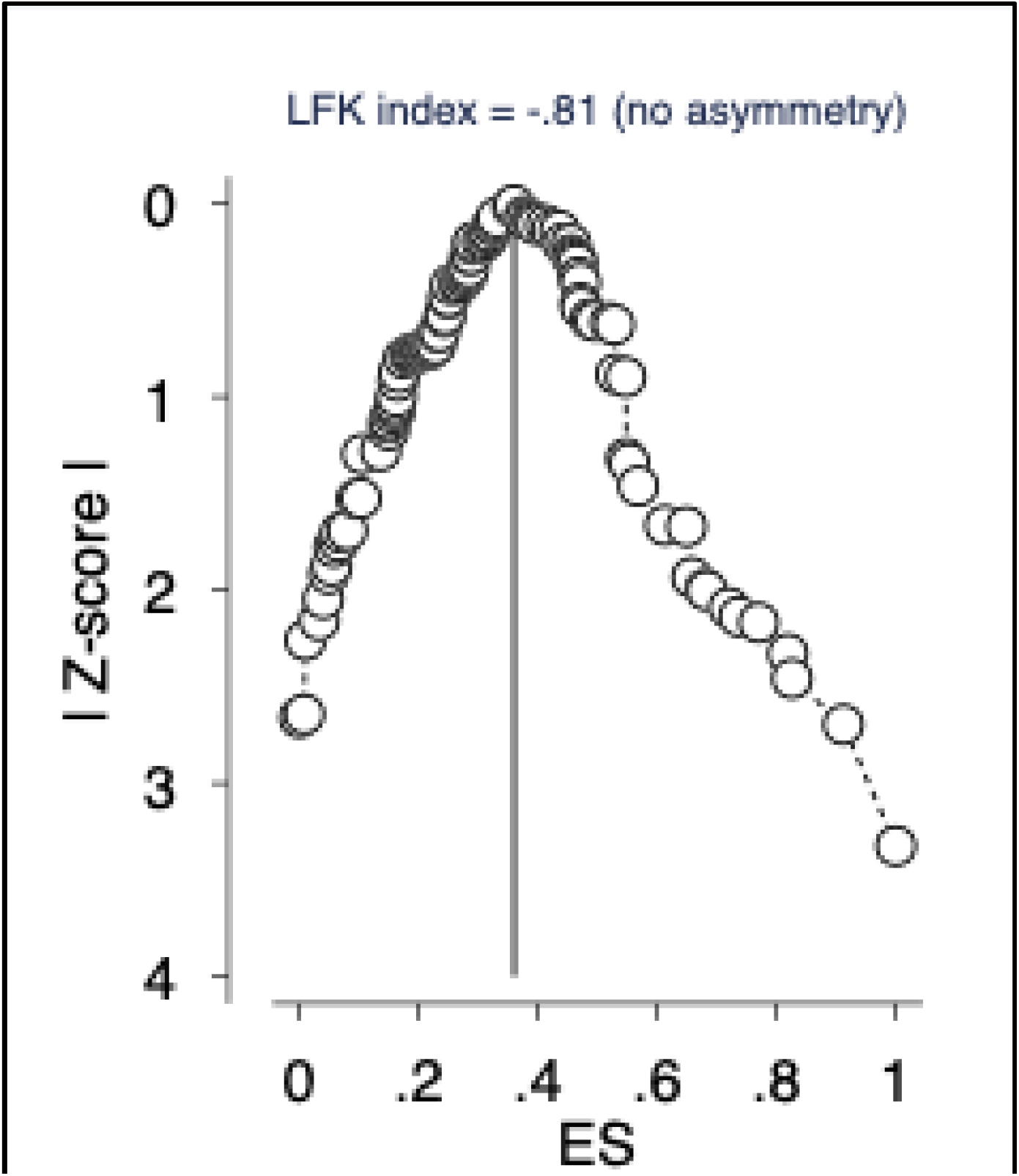
The DOI plot and the Luis Furuya–Kanamori (LFK) index for publication bias Note ES = Effect size.

## Discussion

The analysis of 33 studies with 101,772 participants from Latin America generated pooled prevalence of anxiety, depression, distress, and insomnia of 32%, 27%, 32%, and 35% respectively. Notably, this meta-analysis is the first to investigate the prevalence of mental health symptoms during the COVID-19 crisis in Latin America. The anxiety levels in Latin America were significantly higher than other regions, such as China (25%; p<0.001)(Ren et al., 2020) and Spain (20%; p<0.001)(R. Z. Chen et al., 2021). Latin America has a long-standing history of scarce resources to deal with mental health issues(Alarcón, 2003), which could explain the higher prevalence of mental health symptoms among Latin Americans as revealed by this meta-analysis. Notably, the pooled prevalence of mental health symptoms was lower in Latin America than in Africa and South Asia, as reported by other meta-analyses(J. Chen et al., 2021; Hossain et al., 2020). These cross-region differences may be due to multiple reasons, including heterogeneity in population sizes, variations in and the timing of containment strategies adopted by countries across regions(A.Middelburg & R.Rosendaal, 2020), and the varying degrees of resources available to address mental health issues(Kavita Batra et al., 2020).

The prevalence of mental health symptoms was higher in South America than Central America (33% vs. 27%; p<0.001). This difference might be attributed to variations across these countries in the evolution of the pandemic, the stringency of the COVID-19 responses, and the political climate(Garcia et al., 2020). Previous research noted that South America generally has a high degree of political polarization, which resulted in conflicting information being conveyed to the public which could increase the burden of COVID-19 and its associated psychological corollaries(Garcia et al., 2020). In addition, public health actions or decisions were made mostly at municipal and state levels rather than at central government level, and the lack of central coordination posed several challenges in the control of the pandemic contributing to an increased psychological burden(Garcia et al., 2020).

Based on the evidence of individual studies, our study found a higher prevalence of mental health symptoms among frontline healthcare workers (37%, p<0.001) and university students (36%, p>0.001) than the general population and second-line responders(Batra et al., 2021; Kavita Batra et al., 2020; Min Luo et al., 2020; Pappa et al., 2020). The vulnerabilities of frontline healthcare workers are often attributed to higher risk of infection, burnout, fear of COVID-19 transmission to their family members, and job loss(Bhandari et al., 2021; Xiang et al., 2020). The greater prevalence of mental health symptoms among the university students can be explained by the uncertainties surrounding the course of the pandemic and the sudden transition to online education(Adedoyin & Soykan, 2020; Batra et al., 2021). Moreover, many businesses scaled down their recruitment efforts leaving limited employment for students and more competition in the graduate labour market(Reidy, 2020). These challenges added to the mental health burden among university students.

### Study Limitations

There are a few limitations that merit discussion. First our analysis reveals substantial heterogeneities across studies in the type of survey instruments used and the cut-off scores, both of which may affect the interpretation of the findings. Second, not all Latin American countries have been well-studied, therefore our results may have limited generalizability for the less studied nations. Third, a majority of the included studies were cross-sectional, which provides no information on the prevalence over time during the pandemic. In addition, studies included in this meta-analysis relied on self-reported data of psychological symptoms by the participants and hence do not constitute mental health diagnosis from clinicians. Fourth, other outcomes, such as Post-traumatic Stress Syndrome (PTSD), suicidal ideation, and burnout, were not studied in this meta-analysis, leaving opportunities for prospective studies. Last, a language bias is expected in the study because of the language restriction (only English) applied in this study.

### Practical Implications

First, our systematic review and meta-analysis supports evidence-based medicine by revealing a high proportion of mental health symptoms among the general population and healthcare workers during the COVID-19 pandemic in Latin America. However, our systematic review also reveals there is a lack of evidence in many Latin American countries to guide relevant practice of evidence-based medicine on this topic. Only 9 of the 33 Latin American countries have been studied, leaving 24 countries without any studies to assist the practice of evidence-based healthcare. For instance, no relevant research has been done in Venezuela, the fifth biggest South American country with a population of 28 million, in Chile, the six biggest South American country with a population of 18 million, nor in Guatemala (18 million population), Cuba (11 million population) and Dominican Republic (11 million population), respectively the second, fourth, and fifth most populous countries in Central America. In practice, healthcare organizations in those unstudied countries may use our results in the same region as approximate evidence before direct evidence in those countries emerges.

Our findings that the prevalence of mental health symptoms was higher in South America than Central America (33% vs. 27%; p<0.001) provide evidence to international healthcare organizations, such as the World Psychiatric Association, on their assistance and resource allocation efforts. Our findings of a higher prevalence of mental health symptoms among frontline healthcare workers (37%, p<0.001) and university students (36%, p>0.001) than the general population and general healthcare workers suggest psychiatric and healthcare organizations should prioritize frontline healthcare workers and university students in Latin America.

## Conclusions

This meta-analysis, to our knowledge, provides the first pooled estimates of mental health symptoms among key demographic groups during the COVID-19 crisis in Latin America. The meta-analytical findings of this study underscore the high prevalence of mental health symptoms in Latin Americans during the COVID-19 crisis. Hence, we call for more research to identify people vulnerable to mental health issues to enable evidence-based medicine during the pandemic.

## Data Availability

Not applicable.

## Supplementary materials

**Table S1:**
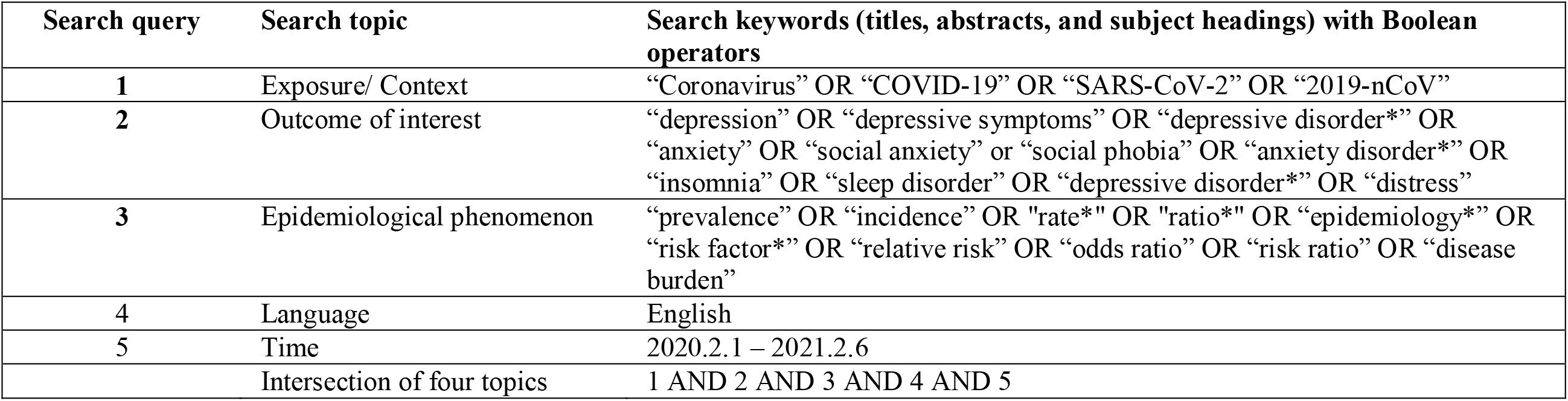
Search strategy of this systematic review and meta-analysis.

| Search query | Search topic | Search keywords (titles, abstracts, and subject headings) with Boolean operators |
| --- | --- | --- |
| 1 | Exposure/ Context | “Coronavirus” OR “COVID-19” OR “SARS-CoV-2” OR “2019-nCoV” |
| 2 | Outcome of interest | “depression” OR “depressive symptoms” OR “depressive disorder*” OR “anxiety” OR “social anxiety” or “social phobia” OR “anxiety disorder*” OR “insomnia” OR “sleep disorder” OR “depressive disorder*” OR “distress” |
| 3 | Epidemiological phenomenon | “prevalence” OR “incidence” OR “rate*” OR “ratio*” OR “epidemiology*” OR “risk factor*” OR “relative risk” OR “odds ratio” OR “risk ratio” OR “disease burden” |
| 4 | Language | English |
| 5 | Time | 2020.2.1 – 2021.2.6 |
|  | Intersection of four topics | 1 AND 2 AND 3 AND 4 AND 5 |

**Table S2:**
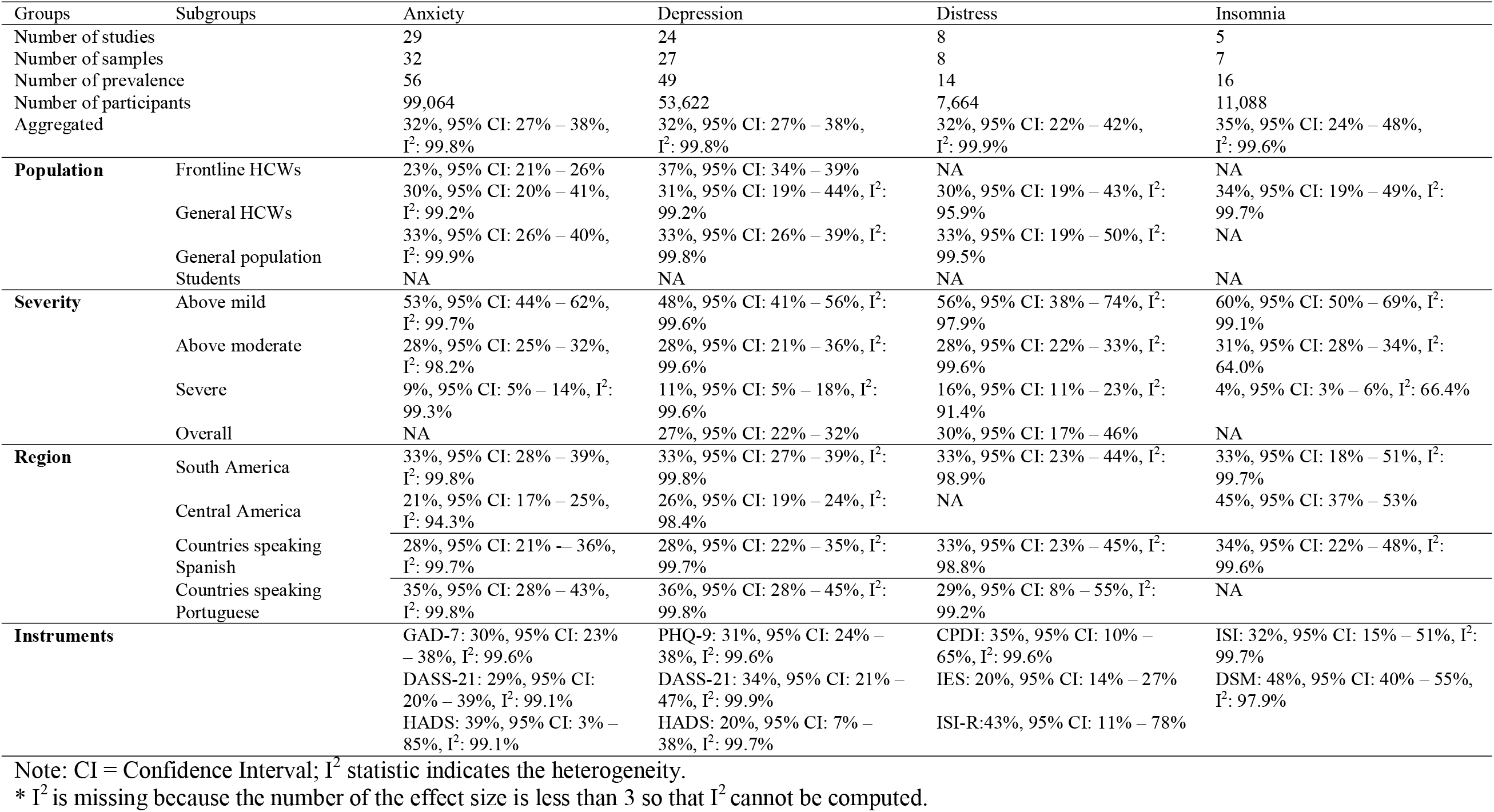
Subgroup analyses of the prevalence of anxiety, depression, and insomnia symptoms.

## Credit author statement

SXZ: Conceptualization, Methodology, Validation, Formal analysis, Investigation, Data curation, Visualization, Writing – original draft, Writing – review & editing, Supervision.

KB: Writing – original draft, Writing – review & editing.

TL: Investigation (Data).

RKD: Investigation (Data).

WX: Investigation (Data).

AY: Investigation (Data).

AD: Investigation (Data), Writing – review & editing.

BZC: Investigation (Data).

RZC: Investigation (Data).

SM: Investigation (Data).

XW: Investigation (Data).

JC: Methodology, Validation, Formal analysis, Investigation, Data curation, Visualization, Writing – original draft, Writing – review & editing, Supervision.

All authors reviewed and approved the manuscript. The corresponding author attests that all listed authors meet authorship criteria and that no others meeting the criteria have been omitted.

## Transparency declaration

The corresponding author affirms this manuscript is an honest, accurate, and transparent account of the study being reported. No important aspects of the study have been omitted and any discrepancies from the study as planned (and, if relevant, registered) have been explained.

## Ethical approval

Not applicable.

## Patient and public involvement

No patient or public was involved in this systematic review and meta-analysis.

## Availability of data

The data that support the findings of this study are available from the corresponding author, J.C., upon reasonable request.

